# Impact of SARS-CoV-2 Vaccines on Covid-19 Incidence and Mortality in the United States

**DOI:** 10.1101/2021.11.16.21266360

**Authors:** Fang Fang, John David Clemens, Zuo-Feng Zhang, Timothy F. Brewer

## Abstract

**Background:** Despite safe and effective vaccines to prevent Severe Acute Respiratory Syndrome coronavirus 2 (SARS-CoV-2) infections and disease, a substantial minority of the US remains resistant to getting vaccinated. It is imperative to know if expanding vaccination rates could reduce community-wide Coronavirus 2019 (COVID-19) disease, not just among those vaccinated.

**Methods:** Negative binomial models were used to estimate associations between U.S. county-level vaccination rates and county-wide COVID-19 incidence and mortality between April 23^rd^ and September 30^th^, 2021. A two-week lag and a four-week lag were introduced to assess vaccination rate impact on incidence and mortality, respectively. Stratified analyses were performed for county vaccination rates 40%, and before and after Delta became the dominant variant.

**Findings:** Among 3,070 counties, each percentage increase in population vaccination rates reduced county-wide COVID-19 incidence by 0·9% (relative risk (RR) 0. 9910 (95% CI: 0·9869, 0·9952)) and mortality by 1·9% (RR 0·9807 (95% CI: 0·9745, 0·9823)). Among counties with vaccination coverage >40%, each percentage increase in vaccination rates reduced COVID-19 disease by 1·5%, RR 0·9850 (95% CI: 0·9793, 0·9952) and mortality by 2·7% (RR 0·9727 (95% CI: 0·9632, 0·9823)). These associations were not observed among counties with <40% vaccination rates. Increasing vaccination rates from 40% to 80% would have reduced COVID-19 cases by 45·4% (RR 0·5458 (95% CI: 0·4335, 0·6873)) and deaths by 67·0% (RR 0·3305 (95% CI: 0·2230, 0·4898)). An estimated 5,989,952 COVID-19 cases could have been prevented and 127,596 lives saved had US population vaccination rates increased from 40% to 80%.

**Interpretations:** Increasing U.S. SARS-CoV-2 vaccination rates results in population-wide reductions in COVID-19 incidence and mortality. Furthermore, increasing vaccination rates above 40% has protective effects among non-vaccinated persons. Given ongoing vaccine hesitancy in the U.S., increasing vaccination rates could better protect the entire community and potentially reach herd immunity.

**Funding:** National Cancer Institute

## INTRODUCTION

Since being recognized in December, 2019, the Severe Acute Respiratory Syndrome coronavirus 2 (SARS-CoV-2) pandemic has caused almost 250 million cases and five million deaths worldwide.^1^ The United States has been particularly affected, with more than 43 million Coronavirus 2019 (COVID-19) cases and 697,000 deaths reported as of September 30^th^, 2021.^2^ Though a number of non-pharmacologic prevention initiatives (NPIs) have been introduced to slow SARS-CoV-2 transmission^3^, vaccines are now recognized as among the most effective means for preventing COVID-19 cases and deaths.^4^

Three vaccine preparations are authorized for use in the US. The BNT162b2 vaccine (Pfizer, Inc. and BioNTech), has full U.S. Food and Drug Administration (FDA) approval^5^, while mRNA-1273 (Moderna) and JNJ-78436735 (Janssen Pharmaceuticals) are available under an emergency use authorization. All three vaccines are effective in preventing SARS-CoV-2 infections and COVID-19 associated disease, hospitalizations, and deaths,^6-8^ though vaccine effectiveness wanes over time and breakthrough infections occur.^4,9^ Vaccine effectiveness also varies by SARS-CoV-2 variant, with higher rates of breakthrough infections or disease reported in vaccinated persons infected with Delta or Beta compared with other variants.^8,10^ With the recognition of waning vaccine immunity and the rise of Delta as the predominant SARS-CoV-2 variant in the U.S., the FDA has authorized the emergency use of a booster for COVID-19 vaccines, including the use of a heterologous booster.^11,12^

Numerous studies have demonstrated that SARS-CoV-2 vaccines are effective in preventing COVID-19 infections and disease in individuals outside of clinical trials, including among health care workers, first responders, individuals attending ambulatory clinics, veterans, and in nursing homes.^13-15^ Despite the proven safety and effectiveness of these vaccines, a substantial minority of the adult US population remains resistant to getting vaccinated.^16^ To understand if the SARS-CoV-2 pandemic can be stopped, it is imperative to understand the impact of vaccination on community-wide SARS-CoV-2 cases and COVID-19 disease, not just among those vaccinated— a concept popularly referred to as “herd immunity”.^17^ To investigate the impact of population percentages of SARS-CoV-2 vaccination on community-wide COVID-19 case and mortality rates, we undertook an ecological analysis of U.S. county vaccination rates on reported county COVID-19 cases and deaths, controlling for socioeconomic, demographic, comorbid conditions, rural/urban, air pollution, hospital capacity, and related factors, the introduction of NPIs and the presence of Delta variant.

## METHODS

Negative binomial models were used to estimate associations between U.S. county-level vaccination rates and county-wide COVID-19 incidence and mortality between April 23^rd^ and September 30^th^, 2021. The dates represent when Delta was first recognized in the U.S. to the end of the study period. To account for the rise of Delta as the dominant U.S. variant, additional analyses were done dividing the study period into before (April 23^rd^ – July 2^nd^) and after (July 3^rd^ – September 30^th^) Delta was responsible for the majority of reported U.S. COVID-19 cases.

Both models were adjusted for the following potential confounders: annual average of ambient atmospheric particulate matter <2·5 µm in diameter (PM_2·5_) between 2000 and 2016, population density, poverty, education, proportions of White, proportions of male, proportion of population older than 65 years old, owner-occupied property, median house value, median household income, percentage of people under 65 years old without health insurance, prevalence of tobacco smoking, and obesity. All covariates were measured at the county level. State-level variables for NPIs policies (facemask mandates, stay home orders) also were included in both models. The mortality model additionally adjusted for the number of county hospital beds.

County-level COVID-19 case and mortality data were obtained from the Johns Hopkins University, Center for Systems Science and Engineering Coronavirus Resource Center (CSSE). CSSE collects county-level confirmed numbers of cases and deaths of 3,342 counties across the U.S. from the U.S. Centers for Disease Control and Prevention (CDC) as well as state departments of health since January 21^st^, 2020.^2^

County-level vaccine data were obtained from Covid Act Now. These data are derived from the U.S. Department of Health and Human Services, the CDC, the New York Times, and official state and county dashboards. Data on vaccinations initiated and vaccination regimens completed were available for all 50 states.^18^ To allow for the development of protective immunity after vaccination, a two-week lag was introduced after people were completely vaccinated (a person vaccinated on June 18^th^ was considered fully protected by July 2^nd^). A two-week lag also was used to account for time between exposure and development of COVID-19 disease. To assess the impact of vaccination on COVID-19 mortality, a four-week lag was used (vaccinated by September 2^nd^ to assess the impact on mortality on September 30^th^).

County-level annual average of PM_2·5_ between the years 2000 and 2016, as well as county-level covariates, were available from a GitHub repository.^19^ County-level socioeconomic and demographic variables for 2018 were available from the US Census/American Community Survey. 2020 data on the prevalence of adult tobacco smoking and adult obesity were accessible through the County Health Rankings & Roadmaps program.^20^ The number of county hospital beds were obtained from an ArcGIS Hub dataset.^21^ State averages were used to replace missing values for county-level prevalence for smoking, obesity, and numbers of hospital beds. U.S. Department of Agriculture 2013 rural and urban codes^22^ were used to identify whether a county was located in metropolitan areas. State-wide non-pharmacologic prevention policies, including facemask use and stay home orders, were obtained from the Boston University School of Public Health.^23^

Counties with invalid Federal Information Processing Standards (n = 10), missing covariates (n = 236), missing vaccination status on June 4^th^ (n = 4), and negative change in incidence (n = 22) or in mortality (n = 61) possibly due to data entry error were excluded. As a result, data from 3,070 counties in 49 states were available to investigate the association between population vaccination rates and county-wide COVID-19 incidence and 3,031 counties in 49 states to investigate the association between vaccination rates and COVID-19 mortality (Table 1).

**Table 1.** Summary of data sources.

| Sources | Description |
| --- | --- |
| Johns Hopkins University Center for Systems Science and Engineering Coronavirus Resource Center (CSSE)<sup>2</sup> | Cumulative county-level confirmed cases and deaths up to September 30, 2021 |
| Covid Act Now<sup>18</sup> | Cumulative county-level number of people completely vaccinated up to September 16, 2021 |
| GitHub repository<sup>19</sup> | Annual average PM<sub>2.5</sub> concentration between 2000 and 2016 |
| The US Census/American Community Survey | County-level socioeconomic and demographic variables in 2018 |
| The County Health Rankings & Roadmaps program<sup>20</sup> | County-level behavioral variables in 2020 |
| ArcGIS hub<sup>21</sup> | Numbers of hospital beds in each county |
| Economic Research Service<sup>22</sup> | Urban/rural code in 2013 |
| Boston University of Public Health<sup>23</sup> | State-level policy of face masks mandates and stay home orders |

As the average US county total population complete vaccination rate at the time of analysis was approximately 40%, further stratified analyses were performed investigating associations between counties where >40% of the total population was completely vaccinated with those with <40% complete population vaccination rates. The impact of 10% incremental increases in total population vaccine coverage was assessed in sensitivity analyses. To account for the correlation within each state, we applied the robust error estimation.^24^ Relative risks (RR) and 95% confidence intervals (CI) are reported. Analyses were performed in SAS 9·4 (Cary, NC). To estimate the number of COVID-19 cases or deaths that could have been prevented during the study period by increasing county-wide vaccination rates, RRs were multiplied by the total at-risk US population. The at-risk population was estimated using the total 2021 U.S. population, subtracting the number of reported COVID-19 cases and deaths when estimating the population at risk for COVID-19 incidence and reported COVID-19 deaths only when estimating the population at risk for COVID-19 mortality.

## RESULTS

Among the 3,070 counties across 49 states, the average county total population complete vaccination rate was 42·3% as of September 16^th^, 2021. Counties with >40% complete vaccination rates had more hospital beds, higher median house values, higher median household incomes, higher population density, and less uninsured population compared with counties with <40% vaccination rates. These counties also were more likely to be located in metropolitan areas and in states where a facemask policy was ever issued before September 30^th^, 2021 (Table 2).

**Table 2.** Characteristics of counties (n = 3,070) in 49 states by vaccination rate as of 9/16.

|  | Total<br>(n = 3,070 in 49<br>states) | Vaccination rate<sup>3</sup><br>on 9/16 ≤ 40%<br>(n = 1,384 in 39<br>states) | Vaccination rate<sup>3</sup><br>on 9/16 > 40%<br>(n = 1,686 in 49<br>states) |
| --- | --- | --- | --- |
|  | Mean (SD) | Mean (SD) | Mean (SD) |
| Incidence of COVID-19 as of 9/30 (%)<sup>1</sup> | 13.75 (3.84) | 14.58 (3.94) | 13.07 (3.63) |
| Incidence of COVID-19 as of 7/2 (%)<sup>1</sup> | 10.25 (3.09) | 10.50 (3.14) | 10.04 (3.04) |
| Mortality of COVID-19 as of 9/30 (%)<sup>2</sup> | 0.24 (0.13) | 0.27 (0.13) | 0.22 (0.12) |
| Mortality of COVID-19 as of 7/2 (%)<sup>2</sup> | 0.21 (0.12) | 0.22 (0.11) | 0.19 (0.11) |
| Full vaccination rate as of 9/16 (%)<sup>3</sup> | 42.27 (10.59) | 33.26 (5.28) | 49.66 (7.74) |
| Full vaccination rate as of 9/2 (%)<sup>3</sup> | 40.60 (10.71) | 31.56 (5.24) | 48.01 (8.03) |
| Full vaccination rate as of 6/18 (%)<sup>3</sup> | 34.75 (10.24) | 26.73 (5.30) | 41.32 (8.48) |
| Full vaccination rate as of 6/4 (%)<sup>3</sup> | 32.95 (9.62) | 25.55 (5.17) | 39.03 (8.05) |
| Average ambient PM<sub>2.5</sub> (µg/m<sup>3</sup>)<sup>4</sup> | 8.41 (2.52) | 8.30 (2.53) | 8.51 (2.51) |
| Ever smokers (%) | 17.45 (3.54) | 18.35 (3.59) | 16.71 (3.32) |
| Adult obesity (%) | 32.88 (5.41) | 33.89 (5.05) | 32.05 (5.55) |
| Number of hospital beds | 398.59 (1103.58) | 175.30 (233.47) | 581.89 (1448.76) |
| Population living in poverty (%) | 15.63 (6.43) | 16.97 (6.24) | 14.53 (6.37) |
| Population density per square mile | 267.33 (1817.39) | 61.91 (171.05) | 435.95 (2434.89) |
| Owner occupied properties (%) | 71.60 (8.06) | 72.61 (7.13) | 70.77 (8.66) |
| Population 25 years old and older with<br>less than high school education (%) | 20.57 (6.09) | 22.48 (5.94) | 19.00 (5.75) |
| White Americans population (%) | 83.51 (16.25) | 84.43 (15.62) | 82.75 (16.73) |
| African Americans population (%) | 9.13 (14.57) | 9.05 (13.95) | 9.20 (15.07) |
| Hispanic population (%) | 9.21 (13.76) | 8.47 (12.08) | 9.82 (14.97) |
| Median house value (×\$1,000) | 145.80 (87.18) | 114.56 (40.55) | 171.45 (105.05) |
| Median household income (×\$1,000) | 51.40 (13.56) | 46.94 (9.94) | 55.07 (14.97) |
| Population over 65 years old (%) | 20.28 (4.81) | 20.25 (4.48) | 20.31 (5.08) |
| Male (%) | 50.06 (2.14) | 50.35 (2.44) | 49.82 (1.82) |
| Uninsured population (%) | 8.88 (3.97) | 10.40 (3.97) | 7.64 (3.52) |
| Metropolitan, n (%) |  |  |  |
| Yes | 1151 (37.49) | 649 (31.43) | 486 (55.16) |
| No | 1919 (62.51) | 1416 (68.57) | 395 (44.84) |
| State stay-home order before 9/30, n (%) |  |  |  |
| Ever issued | 2709 (88.24) | 1218 (88.01) | 1491 (88.43) |
| Never issued | 361 (11.76) | 166 (11.99) | 195 (11.57) |
| State facemask policy before 9/30, n (%) |  |  |  |
| Ever issued | 2297 (74.82) | 867 (62.64) | 1430 (84.82) |
| Never issued | 773 (25.18) | 517 (37.36) | 256 (15.18) |

<sup>1</sup>COVID-19 cases out of total population in each county<sup>2</sup>COVID-19 deaths out of total population in each county<sup>3</sup>Number of people fully vaccinated out of total population in each county<sup>4</sup>Annual average of PM<sub>2.5</sub> between 2000 and 2016

Overall, each percentage increase in a county’s total population vaccination rate between April 23^rd^ and September 30^th^, 2021 was associated with a 0·9% reduction in county-wide COVID-19 cases (relative risk (RR) 0·9910 (95% CI: 0·9869, 0·9952)) and a 1·9% reduction in county-wide COVID-19 mortality (RR 0·9807 (95% CI: 0·9745, 0·9823)). County population vaccination was associated with greater protection against COVID-19 infection, RR of 0·9895 (95% CI: 0·9851, 0·9940), and mortality, RR 0·9742 (95% CI: 0·9670, 0·9804), when the analysis was limited to July 3^rd^ to September 30^th^, corresponding to when Delta became the predominant SARS-CoV-2 variant in the U.S. (Table 3).

**Table 3.**
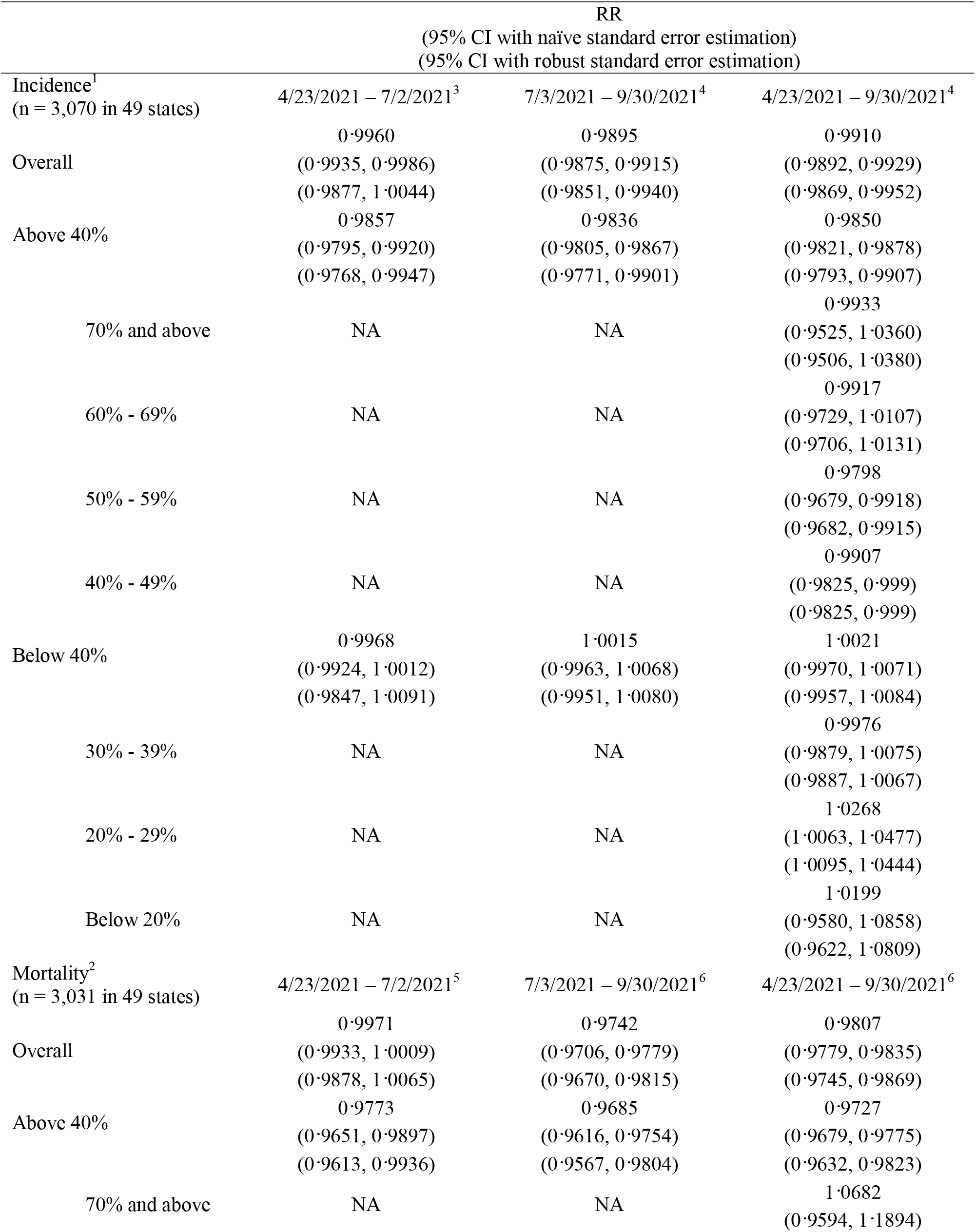
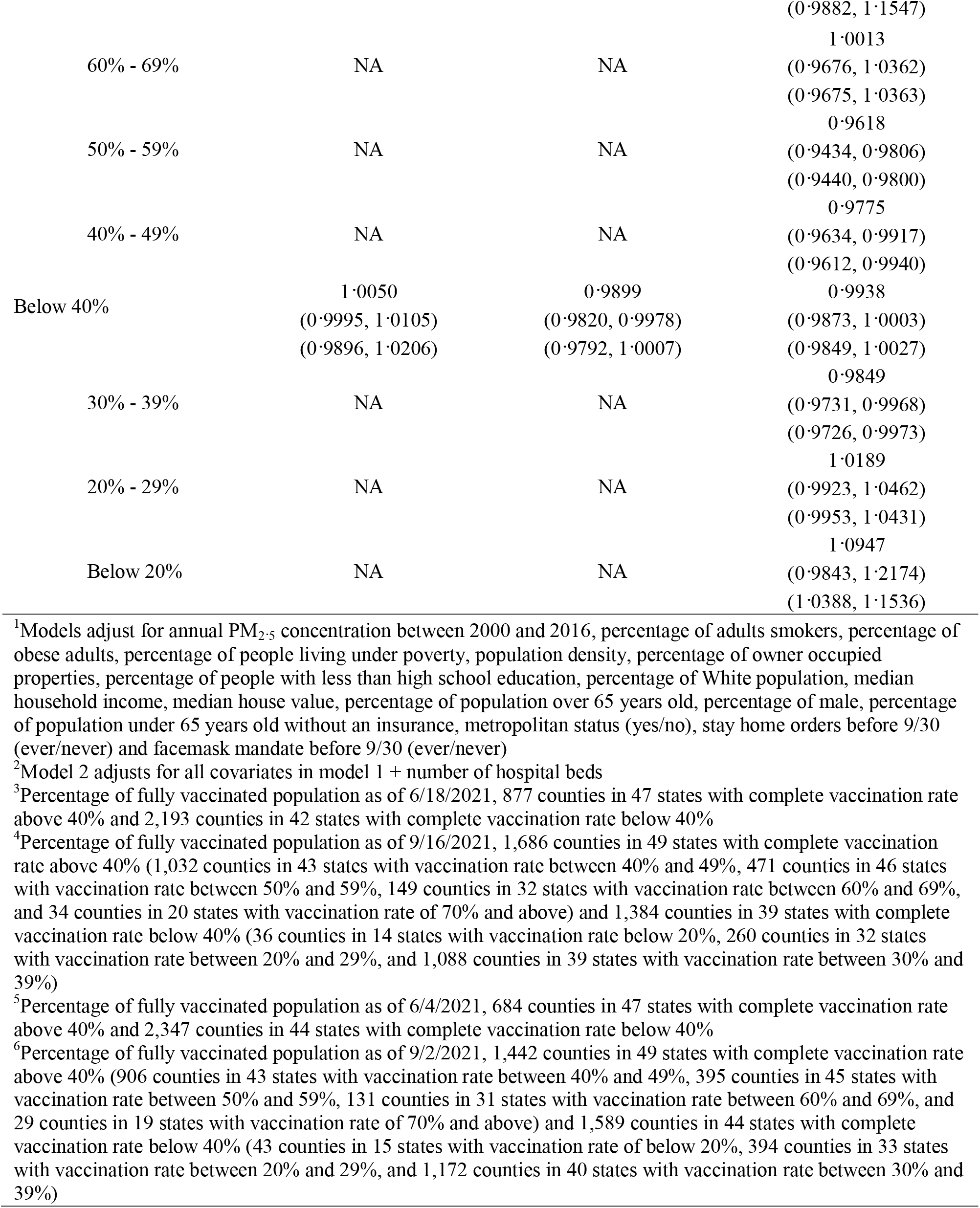
Adjusted RRs of COVID-19 incidence and mortality associated with additional people fully vaccinated per 100 population by vaccination rates.

Among the 1,686 counties in 49 states with complete vaccination coverage >40% as of September 16^th^, each percentage increase in county complete vaccination rates reduced the county population RR of COVID-19 disease by 1·5%, RR 0·9850 (95% CI: 0·9793, 0·9952). However, in counties with <40% complete vaccination rates, the association became null, RR 1·0021 (95% CI: 0·9957, 1·0084). In the 1,442 counties in 49 states with vaccination rate >40% as of September 2^nd^, each percentage increase in the vaccination rate reduced county-wide COVID-19 mortality by 2·7% (RR 0·9727 (95% CI: 0·9632, 0·9823)). In contrast, increasing vaccination rates were not associated with reductions in county-wide COVID-19 mortality among counties with <40% population vaccination rates (RR 0·9938 (95% CI: 0·9849, 1·0027)). Similar results were observed before and after Delta became the most dominant U.S. variant, as well as when further stratifying by 10% incremental increases in vaccination rates (Table 3).

Compared to a county with 40% of its total population being completely vaccinated, increasing the population vaccination rate to 80% would have reduced COVID-19 cases by 45·4% (RR 0·5458 (95% CI: 0·4335, 0·6873)) between April 23^rd^ and September 30^th^ and COVID-19 deaths by 67·0% (RR 0·3305 (95% CI: 0·2230, 0·4898)). Given the at-risk US population, an estimated 5,989,952 COVID-19 cases could have been prevented and 127,596 lives saved had US population vaccination rates increased from 40% to 80% (Table 4 and Table 5).

**Table 4.**
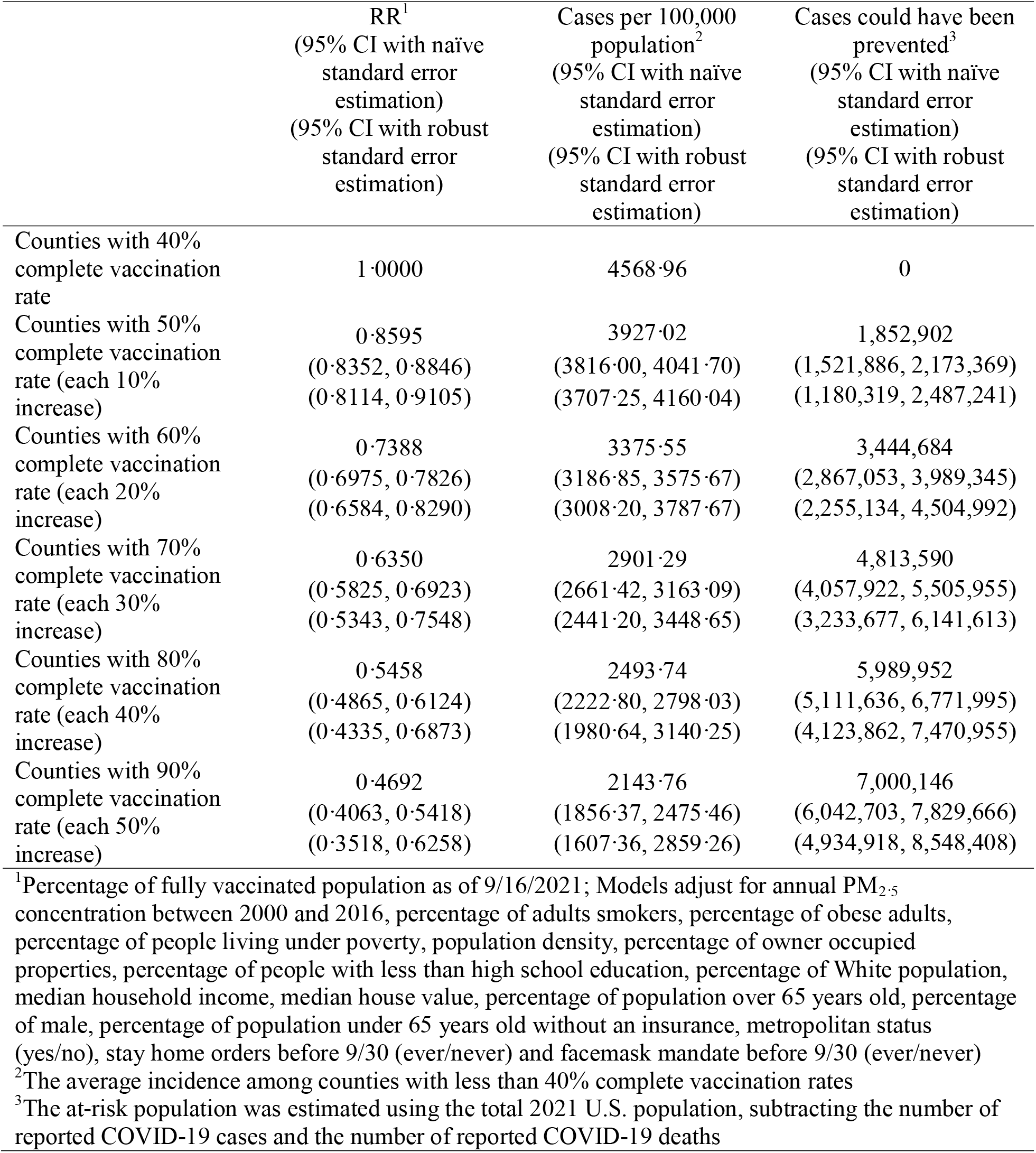
Estimated adjusted RRs, risk reductions of COVID-19 incidence and cases could have been prevented associated with complete vaccination rates between April 23 and September 30, 2021.

**Table 5.**
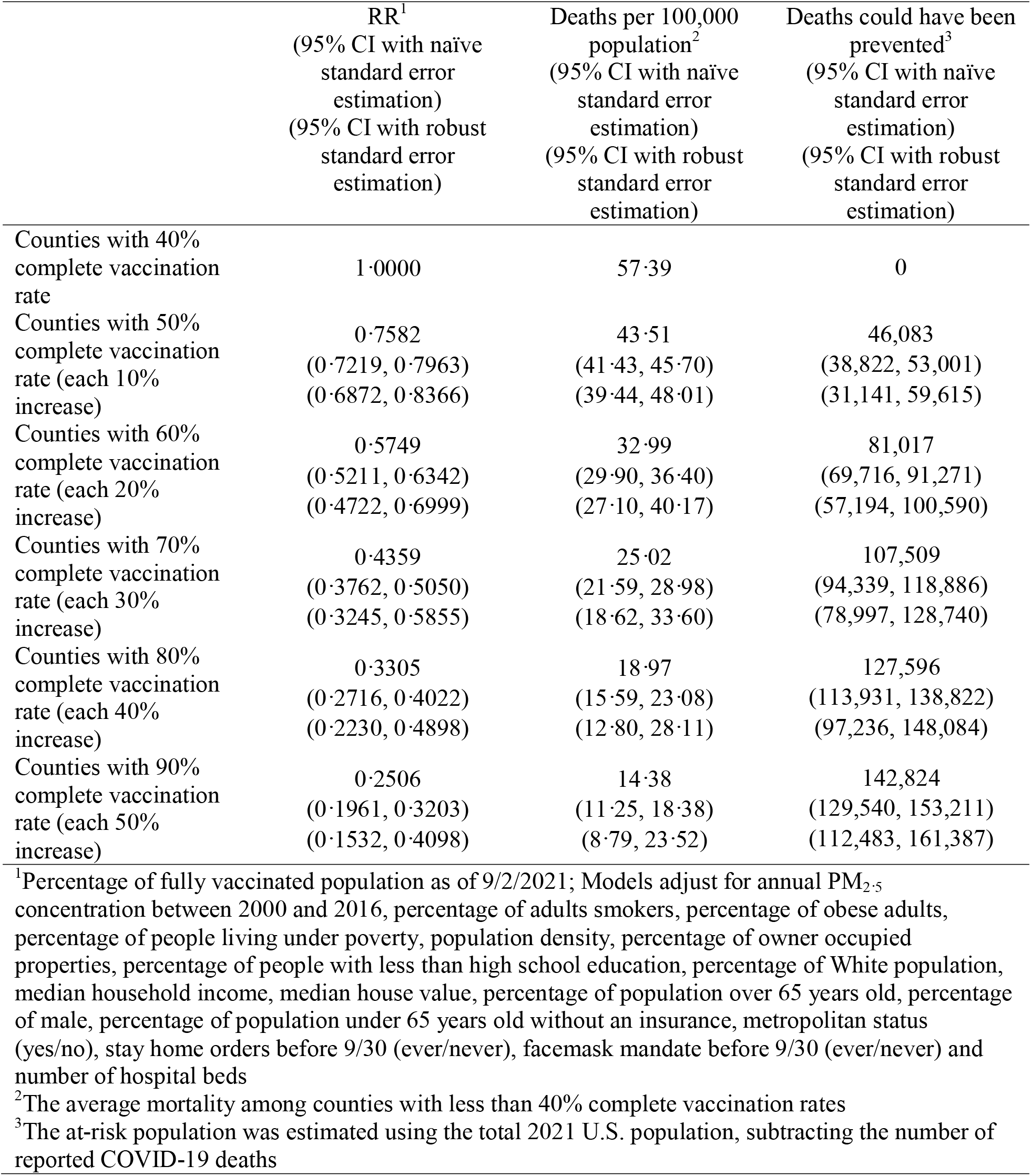
Estimated adjusted RRs, risk reductions of COVID-19 mortality and deaths could have been prevented associated with complete vaccination rates between April 23 and September 30, 2021.

## DISCUSSION

Data from 3,070 counties across 49 states demonstrates that each percentage increase in people being completely vaccinated was associated with a 0·9% decrease in population-wide COVID-19 cases and a 1·9% reduction in COVID-19 mortality between April 23^rd^ and September 30^th^, 2021 after adjusting for potential confounders. Limiting the analysis to between July 3^rd^ and September 30^th^, when Delta was the predominant US variant, gave similar results; each percentage increase in population vaccination rates was associated with a 1·1% decrease in county-wide COVID-19 cases and a 2·6% reduction in COVID-19 deaths. Among the counties with vaccination rate >40%, an additional percentage increase in vaccination rate was associated with a 1·5% reduction in population COVID-19 cases and 2·7% decrease in COVID-19 mortality. In contrast, null associations were observed for counties with <40% coverage of complete vaccination rates. If a county increased its complete vaccination coverage from <40% to 80% during the study period, 45·4% infections and 67·0% deaths due to COVID-19 might have been prevented.

This study is among the first to demonstrate the impact of increasing SARS-CoV-2 vaccination rates on population-wide reductions in COVID-19 incidence and mortality. As the percentage reductions in population COVID-19 cases and deaths generally exceeded the percentage increases in population vaccination rates, these data suggest that increasing population vaccination rates has spillover protective effects among non-vaccinated persons. Moreover, these spillover protective effects grow as population vaccination rates rise. Given ongoing vaccine hesitancy in the US, these results show how increasing vaccination rates could better protect the entire community and potentially reach herd immunity.

We observed clear reductions in COVID-19 cases and deaths with rising population SARS-CoV-2 vaccination rates among counties with >40% vaccination coverage, but not among those with <40% population vaccination rates. This observation was consistent with the Israeli study showing declined COVID-19 incidence as vaccination rates increased.^25^ Similar results were found in the stratified analyses of 10% incremental increases in vaccination rates. Though these data do not enable us to determine the population vaccine rate necessary to achieve herd immunity, the results showed that at least a 40% population vaccine coverage rate is needed to see a protective impact of vaccination among the entire population.

The study was subject to several limitations. First, ecologic study designs are vulnerable to the ecologic fallacy. Therefore, caution is required when interpreting the study results, especially when extrapolating population findings to the individual level. In addition, we cannot rule out the possibility of residual confounding even after controlling for numerous county-level and state-level covariates. Using COVID-19 reported cases may underestimate of the number of actual infections due to under-testing of asymptomatic patients. However, alternative estimates for cumulative incidence, such as seroprevalence^26^, also have limitations including sampling bias, test sensitivity and specificity, and the progress of the pandemic.^27^ Our analysis was not able to assess the impact of the three different vaccines currently available in the U.S., which likely had different efficacies. Although a detailed distribution of different variants in the U.S. was not available, we examined the associations before and after the Delta variant became the predominant strain. Therefore, our results represent the associations between vaccine rates overall and COVID-19 incidence and mortality in the U.S. for vaccines as actually deployed and SARS-CoV-2 variants as they circulated during the period of our analysis.

The differential impact of rising population vaccination rates between before and after the spread of Delta variant might be due to the proportion of counties achieving >40% complete vaccination coverage. Before July 3^rd^, <30% of counties reached 40% population vaccination rates; almost half of counties had >40% of their total population vaccinated by September 30^th^, 2021. Finally, there was limited power to assess impact in counties with vaccination rates <20% or >70%. As a result, counties were combined into either < or >40%, the approximate U.S. county average, to obtain sufficient power and to account for the heterogeneity.

Nevertheless, this study is the first to estimate the association between complete vaccination rates and COVID-19 incidence and mortality in the U.S. general population using county-level data. This nation-wide study covers 3,070 counties in 49 states across the entire country, showing the population-based impact of increasing complete vaccination rates. The proportion of the Delta variant surpassed the proportion of the Alpha variant and accounted for >50% of infection in the U.S. as of June 26^th^, 2021.^12^ Our results indicated that increasing complete vaccination rates were associated with reduced COVID-19 incidence and mortality overall and after July 3^rd^, demonstrating the vaccine impact within communities against SARS-CoV-2 variants in the U.S. including the Delta variant. The population-wide protective associations observed only among counties with >40% vaccination rates urge the uptake of vaccines in all counties. Though we showed estimated COVID-19 incidence and mortality reductions with complete vaccination rates increasing above 40%, these results should be interpreted with caution since there were limited data for counties with very high vaccination coverage rates. Future research should examine the barriers to vaccine uptake, especially among under-privileged populations, as well as estimate the vaccination coverage needed to reach the herd immunity.

## Data Availability

All data produced in the present work are contained in the manuscript

## CONTRIBUTORS

ZF Zhang conceptualized and supervised the project. ZF Zhang and F Fang developed the methods. F Fang obtained the data, conducted the analysis and wrote the original draft. JD Clemens, TF Brewer and ZF Zhang provided feedback on the analyses and edited the draft. All authors reviewed and revised the manuscript for critical content.

## ACKNOWLEDGEMENTS

We acknowledged all organizations and groups collecting and maintaining the publicly available data sources listed in Table 1 and utilized in this study. F Fang is supported by the NIH/NCI T32 CA009142. The funding source did not involve in this study.

